# Geospatial Analysis of Antenatal Care Utilization and Its Determinants Among Women in Ghana: Evidence from 2022 Demographic and Health Survey

**DOI:** 10.64898/2026.05.27.26354191

**Authors:** Samuel Yaw Opoku, Enoch Weikem Weyori, Sabina Ampon-Wireko, Mathius Justice Akanzabwon Asaarik, Felix Fiavor, Theresah Owusua, Patience Nawaane

**Affiliations:** Catholic University of Ghana, Department of Public Health, Fiapre-Sunyani; Nursing and Midwifery Training College, Sunyani

**Keywords:** Antenatal care utilization, Maternal healthcare, Geographic inequalities

## Abstract

**Background:** Antenatal care (ANC) utilization is critical for improving maternal and neonatal health outcomes. Despite the World Health Organization’s recommendation of at least eight ANC contacts during pregnancy and the implementation of free maternal healthcare policies in Ghana, significant geographic and socioeconomic disparities in ANC utilization persist. This study therefore assessed the spatial distribution and geographically varying determinants of adequate ANC utilization among women in Ghana.

**Methods:** A cross-sectional analytical study was conducted using women’s data from the 2022 Ghana Demographic and Health Survey. The analysis included women aged 15–49 years with an index child younger than five years preceding the survey. Descriptive statistics were computed using Stata version 18, while spatial analyses were conducted in QGIS version 3.44. Global Moran’s I was used to assess spatial autocorrelation, whereas Local Moran’s I and Getis-Ord Gi* analyses identified spatial clusters, hotspots, and coldspots of ANC utilization. Ordinary Least Squares (OLS) regression and Geographically Weighted Regression (GWR) models were fitted to assess global and local determinants of ANC utilization.

**Results:** Overall, only 26.0% of women achieved adequate ANC utilization, while 74.0% reported inadequate ANC attendance. Adequate ANC utilization was higher among women with higher education (42.0%) and those from the richest households (41.3%) compared with women without formal education (19.1%) and those from the poorest households (17.6%). Regional disparities were observed, with Western (48.8%), Eastern (48.0%), and Greater Accra (47.3%) regions recording the highest ANC utilization, whereas Savannah (24.7%), Northern (25.8%), and North East (26.8%) regions recorded the lowest utilization levels. Global Moran’s I demonstrated significant positive spatial autocorrelation (Moran’s I = 0.457, p = 0.044), indicating geographic clustering of ANC utilization across Ghana. Getis-Ord Gi* analysis identified significant coldspots within Northern, Savannah, and North East regions, while Central Region demonstrated significant hotspot clustering. OLS regression showed that maternal education (β = 0.284, p = 0.003) and household wealth (β = 0.191, p = 0.011) positively influenced ANC utilization, whereas distance to health facility negatively influenced utilization (β = -0.156, p = 0.019). The GWR model demonstrated improved explanatory performance (Adjusted R² = 0.71), confirming substantial spatial heterogeneity in ANC determinants across Ghana.

**Conclusion:** Adequate ANC utilization in Ghana remains low and geographically unequal. Maternal education, household wealth, and geographic accessibility significantly influence ANC utilization, with pronounced disparities concentrated within Northern Ghana. Spatially targeted maternal health interventions aimed at improving education, reducing socioeconomic inequalities, and enhancing healthcare accessibility are required to improve equitable ANC utilization across Ghana.

## Introduction

Maternal mortality remains a major public health concern globally, particularly in low- and middle-income countries where access to quality maternal healthcare services remains inadequate. Antenatal care (ANC) is a critical component of maternal healthcare that contributes substantially to the prevention, early detection, and management of pregnancy-related complications, thereby improving maternal and neonatal outcomes (Chamani et al., 2021; Kimario et al., 2025; Tilahun et al., 2026). In response to persistent maternal mortality, the World Health Organization (WHO) revised its ANC model in 2016 and recommended a minimum of eight ANC contacts during pregnancy to enhance positive pregnancy experiences and reduce adverse maternal and neonatal outcomes (Akum et al., 2023; Lee et al., 2024; Owusu et al., 2026). Despite these recommendations, utilization of adequate ANC services remains suboptimal across many countries in sub-Saharan Africa (Amponsah-Tabi et al., 2022; Chamani et al., 2021).

In Ghana, maternal healthcare utilization has improved over the past decade following the implementation of several maternal health interventions, including the free maternal healthcare policy under the National Health Insurance Scheme (Adawudu et al., 2024). Nevertheless, substantial inequalities in ANC utilization continue to exist across socioeconomic and geographic populations (Raphael, 2025). Recent findings from the Ghana Demographic and Health Survey indicated that less than half of women achieved the recommended eight or more ANC visits during pregnancy, with marked disparities observed across regions (Aboagye et al., 2024a). Southern regions such as Greater Accra, Eastern, and Western generally report higher ANC utilization, whereas Northern, Savannah, North East, and Oti regions continue to demonstrate lower utilization levels (Aboagye et al., 2024a). These disparities suggest the presence of important socioeconomic and geographic barriers affecting maternal healthcare utilization across the country.

Previous studies in Ghana and other sub-Saharan African countries have identified maternal education, household wealth, parity, health insurance status, and distance to health facilities as important determinants of ANC utilization (Hajjar et al., 2026; Zitierung et al., 2025). Women with higher educational attainment and better socioeconomic status are more likely to utilize maternal healthcare services adequately, while women residing in rural and underserved communities often experience substantial financial and geographic barriers to accessing care. Geographic accessibility remains particularly important in Ghana, where unequal distribution of healthcare facilities and transportation challenges continue to affect healthcare utilization among vulnerable populations (Agbenyo et al., 2017; Anumudu et al., 2025; Kota et al., 2023; Zitierung et al., 2025).

Although several studies have examined determinants of ANC utilization in Ghana using conventional statistical approaches, limited attention has been given to the spatial distribution and geographic clustering of ANC utilization across the country (Aboagye et al., 2024b; Belay et al., 2025). Maternal healthcare utilization is inherently spatial because neighboring regions often share similar socioeconomic conditions, healthcare infrastructure, cultural practices, and accessibility patterns (Belay et al., 2025). Conventional regression models may therefore fail to adequately capture spatial dependence and geographic heterogeneity in ANC utilization patterns (Tareke et al., 2022). Spatial analytical approaches such as Moran’s I, Getis-Ord Gi*, and Geographically Weighted Regression (GWR) provide important opportunities to identify hotspots, coldspots, and geographically varying determinants of ANC utilization that may not be detected using traditional analytical methods (Belay et al., 2025; Tareke et al., 2022).

Understanding the spatial distribution and determinants of ANC utilization is important for developing geographically targeted maternal health interventions and reducing regional inequalities in maternal healthcare utilization across Ghana. However, evidence on the spatial heterogeneity and geographically varying determinants of adequate ANC utilization in Ghana remains limited. Therefore, this study aimed to assess the spatial distribution, clustering patterns, and geographically varying socio-economic determinants of adequate ANC utilization among women with children under five years using data from the Ghana Demographic and Health Survey.

## Methodology

### Study Design and Data Source

This study employed a cross-sectional analytical design using secondary data obtained from the Demographic and Health Surveys Program Demographic and Health Survey (DHS). The DHS is a nationally representative household survey conducted periodically to provide reliable estimates on population health, maternal and child health, fertility, and healthcare utilization indicators in low- and middle-income countries.

The analysis utilized women’s recode data extracted from the most recent Ghana DHS dataset. The DHS applies a standardized methodology across participating countries, thereby ensuring comparability, reliability, and validity of survey estimates. Data collection was implemented using structured questionnaires administered to eligible women of reproductive age (15–49 years).

### Study Population

The study population comprised women aged 15–49 years who had a live birth within five years preceding the survey. The analysis was restricted to women whose most recent or index child was less than five years old at the time of the survey to minimize recall bias regarding antenatal care utilization during pregnancy.

Women with incomplete information on antenatal care attendance or missing geographic coordinates were excluded from the spatial analyses. After applying inclusion and exclusion criteria, a weighted sample of eligible women was included in the final analysis.

### Sampling Procedure

The DHS employed a stratified two-stage cluster sampling design to obtain nationally representative estimates. In the first stage, enumeration areas (EAs) were selected from the national sampling frame developed from the most recent population and housing census using probability proportional to size sampling techniques. In the second stage, households were systematically selected within each sampled cluster.

Sampling weights provided within the DHS dataset were applied during the analysis to account for unequal probabilities of selection, clustering, and stratification effects inherent in the survey design. The weighting procedure ensured national representativeness of the findings.

### Study Variables

#### Outcome Variable

The primary outcome variable was adequate antenatal care (ANC) utilization. ANC utilization was operationalized using the World Health Organization (WHO) 2016 recommendation of eight or more ANC visits during pregnancy.

Women who attended: fewer than eight ANC visits were categorized as having inadequate ANC utilization, while women who attended eight or more ANC visits were categorized as having adequate ANC utilization.

The outcome variable was coded as: Inadequate ANC utilization (<8 visits) coded as 0 and Adequate ANC utilization (≥8 visits) as 1.

#### Explanatory Variables

The explanatory variables included socioeconomic, demographic, and geographic characteristics identified from previous literature as determinants of maternal healthcare utilization. These variables included: Maternal education level, Household wealth index, Distance to health facility, and Count of pregnant women/population burden.

- Maternal education was categorized into: No formal education, Primary education, Secondary education, and Higher education
- Household wealth index was categorized into: Poorest, Poorer, Middle, Richer, and Richest
- Distance to health facility was categorized as: Less than 5 km and Greater than 5 km.

### Data Management and Statistical Analysis

Data management and statistical analyses were performed using Stata version 19.0 and QGIS version 3.44. Sampling weights, clustering, and stratification variables were incorporated using the DHS survey design approach to account for the complex sampling structure. Descriptive statistics including frequencies, percentages, proportions, and confidence intervals were computed to summarize respondent characteristics and ANC utilization patterns across Ghana.

Regional prevalence of adequate ANC utilization was calculated and spatially aggregated at the regional level for spatial analyses.

### Spatial Analysis

Spatial analyses were conducted to assess the geographic distribution and spatial dependence of ANC utilization across Ghana. Geographic coordinates of DHS clusters were linked to regional administrative boundaries using spatial join procedures in QGIS.

To preserve respondent confidentiality, DHS cluster coordinates were geographically displaced in accordance with DHS data protection protocols. Urban clusters were displaced up to 2 km, while rural clusters were displaced up to 5 km, with a small proportion displaced up to 10 km.

### Global Spatial Autocorrelation

Global spatial autocorrelation of ANC utilization was assessed using the Moran’s I statistic to determine whether ANC utilization patterns were clustered, dispersed, or randomly distributed across Ghana.

A positive Moran’s I value indicated clustering of similar ANC utilization patterns, whereas negative values indicated spatial dispersion. Statistical significance was determined using corresponding z-scores and p-values at a 95% confidence level.

### Local Spatial Cluster Analysis

Local spatial clustering of ANC utilization was assessed using the Local Moran’s I analysis, and Getis-Ord Gi* hotspot analysis.

Local Moran’s I analysis identified High-High (HH), Low-Low (LL), High-Low (HL), and Low-High (LH) as spatial clusters of ANC utilization across Ghana whiles Getis-Ord Gi* hotspot analysis was used to identify statistically significant hotspots and coldspots of adequate ANC utilization. Regions with positive z-scores represented hotspots, while regions with negative z-scores represented coldspots. Statistical significance was assessed at 90%, 95%, and 99% confidence levels.

### Bivariate Spatial Autocorrelation Analysis

Bivariate Local Moran’s I analysis was conducted to assess the spatial relationships between ANC utilization and selected socioeconomic determinants including maternal education, household wealth index, and distance to health facility.

This analysis identified regions where high or low ANC utilization spatially corresponded with similar or contrasting socioeconomic characteristics across neighboring regions.

### Ordinary Least Squares Regression

An Ordinary Least Squares Regression model was initially fitted to examine the global relationship between ANC utilization and selected explanatory variables. The OLS model served as the baseline regression model prior to spatial regression analysis.

Model diagnostics including adjusted R², Akaike Information Criterion (AIC), variance inflation factors (VIF), and residual Moran’s I were assessed to evaluate model performance, multicollinearity, and residual spatial autocorrelation.

### Geographically Weighted Regression

Accounting for spatial non-stationarity in ANC utilization determinants across Ghana, Geographically Weighted Regression analysis was conducted.

An adaptive Gaussian kernel was applied, while bandwidth selection was optimized using the corrected Akaike Information Criterion (AICc). GWR generated local regression coefficients for each region, allowing examination of spatially varying relationships between ANC utilization and explanatory variables. Model performance was evaluated using Residual Sum of Squares (RSS), sigma values, adjusted R², AIC, and AICc statistics.

### Ethical Considerations

The study utilized publicly available secondary DHS data obtained with permission from the DHS Program. The DHS surveys received ethical approval from relevant national ethics committees and institutional review boards prior to data collection. All respondents provided informed consent before participation.

No personally identifiable information was included in the dataset used for analysis, and confidentiality of respondents was maintained throughout the study.

## Findings and Results

### 3.1 Socio-demographic characteristics

The findings demonstrated that adequate ANC utilization increased with higher educational attainment, ranging from 19.1% among women with no formal education to 42.0% among women with higher education. Similarly, women from the richest households recorded higher adequate ANC utilization (41.3%) compared with those from the poorest households (17.6%).

Women residing within 5 km of a health facility demonstrated higher adequate ANC utilization (28.4%) than those living more than 5 km away (20.1%), suggesting that geographic accessibility influences maternal healthcare utilization.

Regional disparities were also observed across Ghana. Western (35.0%), Eastern (32.6%), Upper East (32.0%), and Greater Accra (31.9%) regions recorded the highest adequate ANC utilization, whereas Savannah (19.2%), Oti (19.7%), Northern (20.5%), and Western North (21.3%) regions recorded the lowest utilization levels.

**Table 1:** Socio-demographic characteristics of respondents.

| Variable | ANC utilization in pregnancy<br>Weighted |  | Total<br>[N=6965] |
| --- | --- | --- | --- |
|  | Not adequate<br>[n=2442] | Adequate<br>[n=4523] |  |
| Education |  |  |  |
| No Education | 1666 (80.9) | 394 (19.1) | 2060 (29.6) |
| Primary | 882 (78.3) | 244 (21.7) | 1216 (16.2) |
| Secondary | 2299 (70.8) | 949 (29.2) | 3248 (46.6) |
| Higher | 308 (58.0) | 223 (42.0) | 531 (7.6) |
| **Wealth index** |  |  |  |
| Poorest | 1760 (82.4) | 376 (17.6) | 2136 (30.7) |
| Poorer | 1264 (74.8) | 425 (25.2) | 1689 (24.3) |
| Middle | 941 (72.8) | 351 (27.2) | 1292 (18.6) |
| Richer | 722 (68.7) | 329 (31.3) | 1051 (15.1) |
| Richest | 468 (58.7) | 329 (41.3) | 797 (11.4) |
| **Distance to health facility** |  |  |  |
| < 5 km | 3547 (71.6) | 1406 (28.4) | 4953 (71.1) |
| > 5 km | 1608 (79.9) | 404 (20.1) | 2012 (28.9) |
| **Regional distribution** |  |  |  |
| Western | 219 (65.0) | 118 (35.0) | 337 (4.8) |
| Central | 274 (71.5) | 109 (28.5) | 383 (5.5) |
| Greater Accra | 246 (68.1) | 115 (31.9) | 361 (5.2) |
| Volta | 215 (71.9) | 84 (28.1) | 299 (4.3) |
| Eastern | 225 (67.3) | 109 (32.6) | 334 (4.8) |
| Ashanti | 329 (71.2) | 133 (28.8) | 462 (6.6) |
| Western North | 225 (78.7) | 69 (21.3) | 324 (4.7) |
| Ahafo | 265 (69.9) | 114 (30.1) | 379 (5.4) |
| Bono | 232 (71.6) | 92 (28.4) | 324 (4.7) |
| Bono East | 365 (74.8) | 123 (25.2) | 488 (7.0) |
| Oti | 379 (80.3) | 93 (19.7) | 472 (6.8) |
| Northern | 545 (79.5) | 141 (20.5) | 686 (9.9) |
| Savannah | 458 (80.8) | 109 (19.2) | 567 (8.1) |
| North East | 476 (78.2) | 133 (21.8) | 609 (8.7) |
| Upper East | 327 (68.0) | 154 (32.0) | 481 (6.9) |
| Upper West | 345 (75.2) | 114 (24.8) | 459 (6.6) |

### 3.2 Prevalence of ANC utilization

The pie chart illustrates the prevalence of antenatal care (ANC) attendance among the study population. The majority of respondents reported inadequate ANC attendance (74.0%), while only 26.0% achieved adequate ANC attendance.

**Figure 1:**
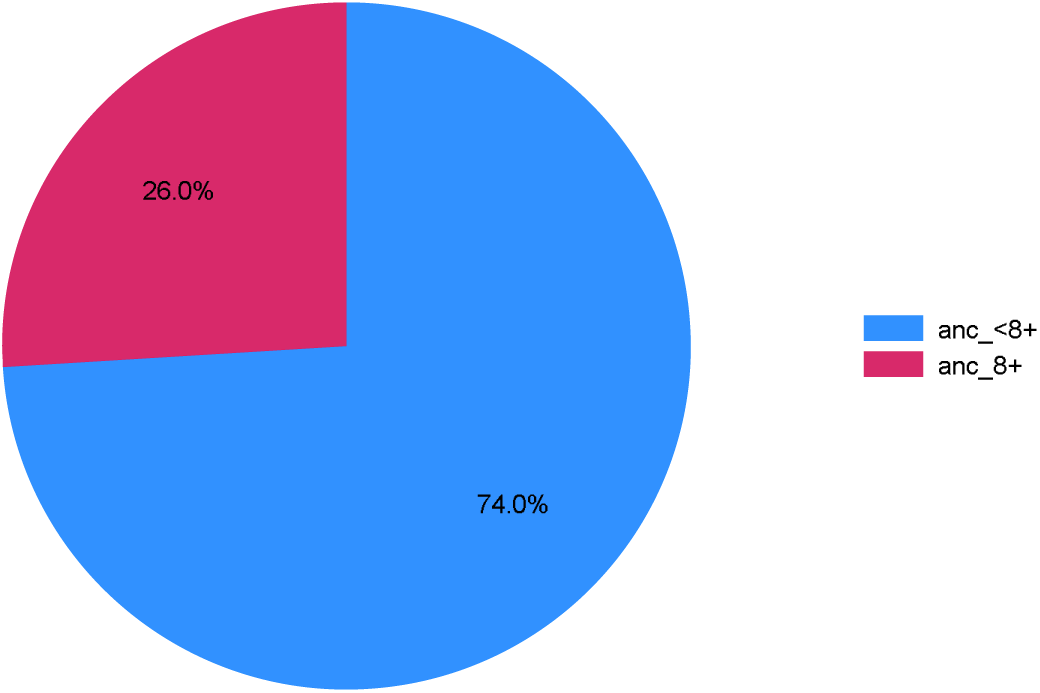
Prevalence of ANC utilization

### 3.3 Regional distribution of ANC utilization

The findings demonstrated substantial regional disparities in ANC 8+ utilization across Ghana, with Western (48.8%), Eastern (48.0%), and Greater Accra (47.3%) regions recording the highest ANC utilization, while Savannah (24.7%), Northern (25.8%), and North East (26.8%) regions recorded the lowest utilization levels. Regions with higher educational attainment and wealth indices generally exhibited better ANC utilization patterns, whereas northern regions characterized by lower socioeconomic indicators and longer distances to health facilities demonstrated poorer ANC attendance. These findings highlight the influence of socioeconomic and geographic inequalities on maternal healthcare utilization across Ghana.

**Table 2:**
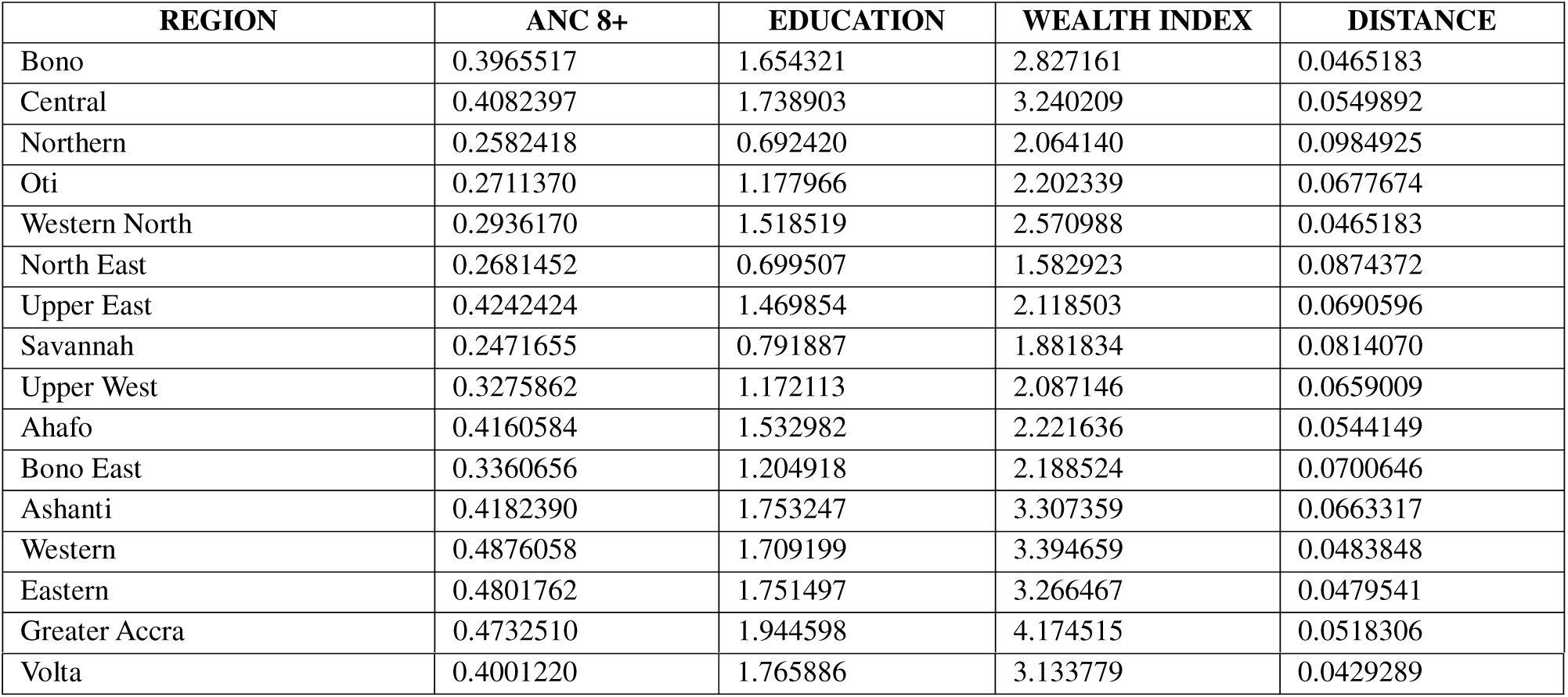
Regional distribution of ANC utilization in Ghana.

| REGION | ANC 8+ | EDUCATION | WEALTH INDEX | DISTANCE |
| --- | --- | --- | --- | --- |
| Bono | 0.3965517 | 1.654321 | 2.827161 | 0.0465183 |
| Central | 0.4082397 | 1.738903 | 3.240209 | 0.0549892 |
| Northern | 0.2582418 | 0.692420 | 2.064140 | 0.0984925 |
| Oti | 0.2711370 | 1.177966 | 2.202339 | 0.0677674 |
| Western North | 0.2936170 | 1.518519 | 2.570988 | 0.0465183 |
| North East | 0.2681452 | 0.699507 | 1.582923 | 0.0874372 |
| Upper East | 0.4242424 | 1.469854 | 2.118503 | 0.0690596 |
| Savannah | 0.2471655 | 0.791887 | 1.881834 | 0.0814070 |
| Upper West | 0.3275862 | 1.172113 | 2.087146 | 0.0659009 |
| Ahafo | 0.4160584 | 1.532982 | 2.221636 | 0.0544149 |
| Bono East | 0.3360656 | 1.204918 | 2.188524 | 0.0700646 |
| Ashanti | 0.4182390 | 1.753247 | 3.307359 | 0.0663317 |
| Western | 0.4876058 | 1.709199 | 3.394659 | 0.0483848 |
| Eastern | 0.4801762 | 1.751497 | 3.266467 | 0.0479541 |
| Greater Accra | 0.4732510 | 1.944598 | 4.174515 | 0.0518306 |
| Volta | 0.4001220 | 1.765886 | 3.133779 | 0.0429289 |

### 3.4 Global Moran’s I spatial autocorrelation

The Moran’s I analysis demonstrated statistically significant positive spatial autocorrelation in ANC utilization across Ghana (Moran’s I = 0.457, p = 0.044). The observed Moran’s I value exceeded the expected Moran’s I under spatial randomness (Expected I = -0.067), indicating significant geographic clustering of similar ANC utilization patterns across neighboring regions. The standard deviation of 0.343 further reflects variability in the spatial distribution of ANC utilization across the study area.

The statistically significant p-value (p = 0.044) suggests that the observed spatial pattern was unlikely to have occurred by chance. Overall, the findings indicate that neighboring regions tended to exhibit dissimilar ANC utilization patterns, suggesting the presence of spatial heterogeneity and regional inequalities in maternal healthcare utilization across Ghana.

**Figure 2:**
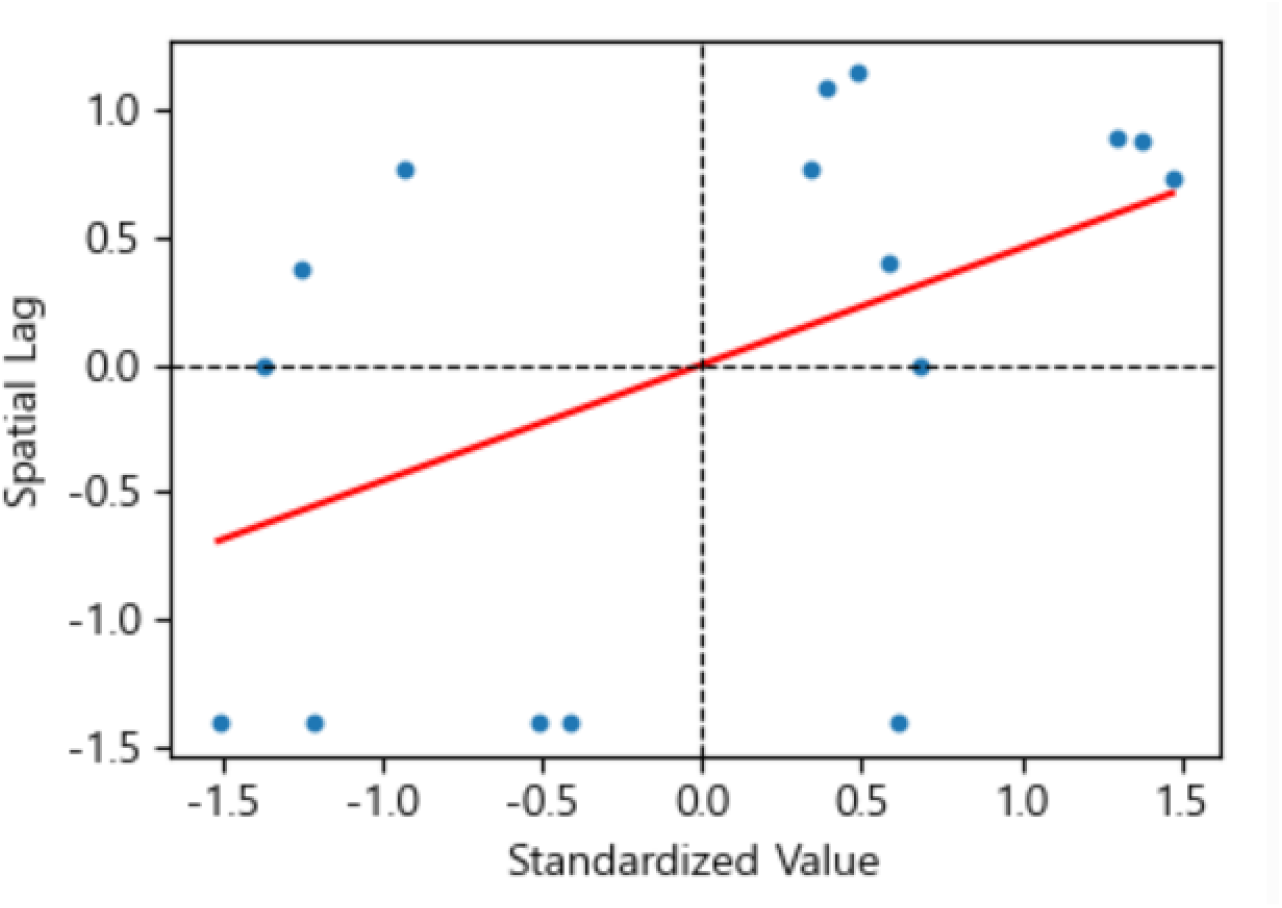
Moran’s I spatial autoregression [**Moran’s I**: 0.45702 | **Expectation**: -0.06667 | **Std Dev**: 0.35693 | **p-value**: 0.0437]

### 3.5 Local Moran’s I cluster analysis

The Local Moran’s I analysis revealed significant spatial clustering of ANC utilization across Ghana, demonstrating substantial regional inequalities in maternal healthcare utilization. Strong clustering patterns were predominantly observed within northern Ghana, particularly in Northern (ANC = 0.258; Moran’s I = 2.982; p = 0.010), North East (ANC = 0.268; Moran’s I = 1.528; p = 0.004), Savannah (ANC = 0.247; Moran’s I = 0.799; p = 0.031), and Upper West (ANC = 0.328; Moran’s I = 0.216; p = 0.025) regions, where lower ANC utilization levels were geographically concentrated among neighboring regions. Conversely, southern regions including Western (ANC = 0.488; Moran’s I = 0.627), Eastern (ANC = 0.480; Moran’s I = 0.093), and Greater Accra (ANC = 0.473; Moran’s I = 0.445) recorded relatively higher ANC utilization with comparatively weaker clustering intensity. Overall, the findings indicate significant spatial dependence in ANC utilization across Ghana and suggest that geographic and regional contextual factors substantially influence maternal healthcare utilization patterns.

**Figure 3:**
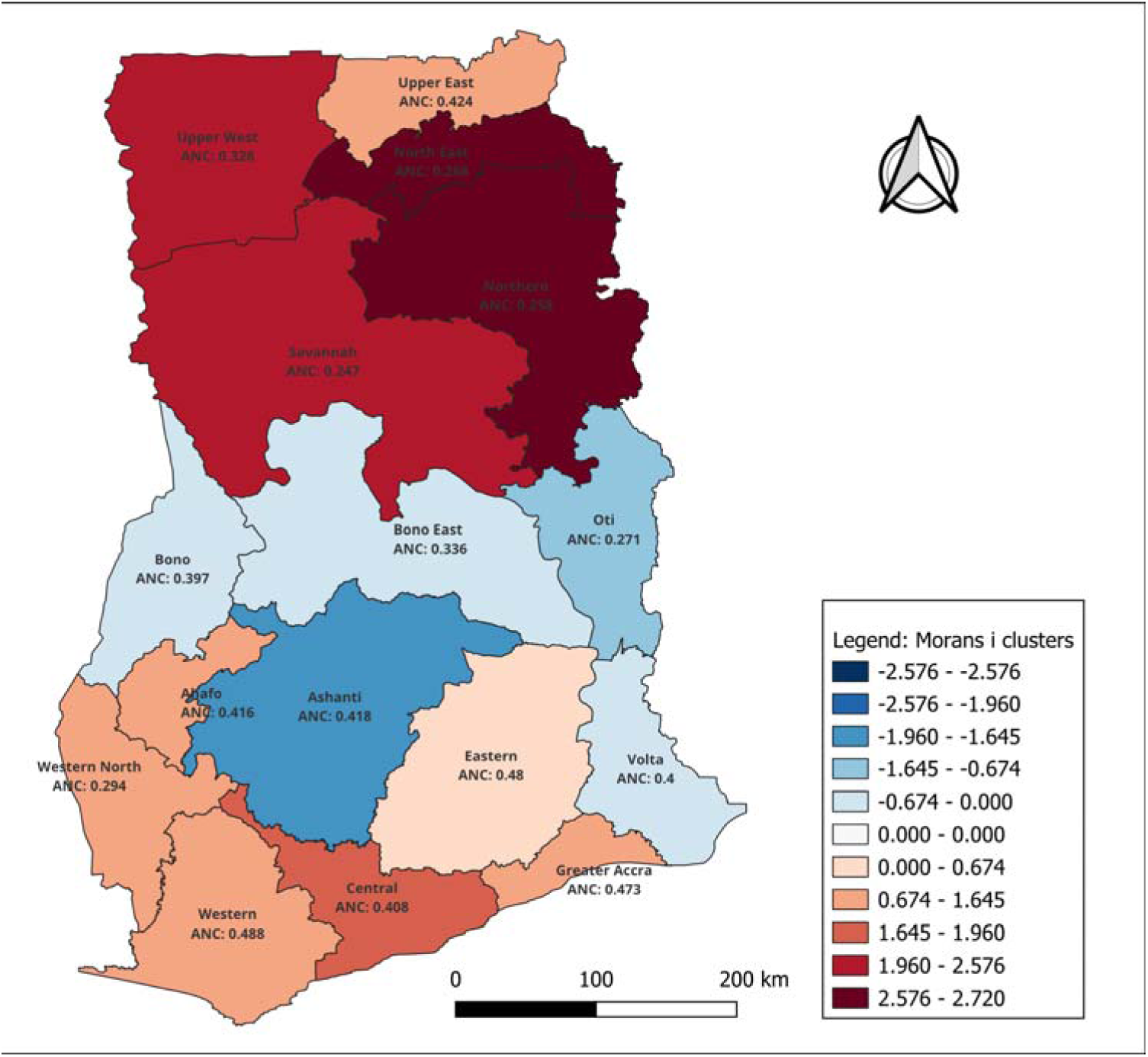
Local Moran’s I Cluster Map of Adequate ANC Utilization Across Ghana

### 3.6 Getis-Ord Gi* hotspot analysis

Getis-Ord Gi* hotspot analysis revealed significant spatial disparities in adequate antenatal care (ANC) utilization across Ghana. Adequate ANC utilization, defined as attendance of eight or more ANC visits, demonstrated significant coldspot clustering within northern Ghana. Northern Region (ANC = 0.26) exhibited a 99% confidence coldspot, while Savannah (ANC = 0.25) and North East (ANC = 0.27) regions demonstrated 95% confidence coldspots, indicating significant geographic concentration of inadequate ANC utilization within these areas.

Conversely, significant hotspot clustering of relatively higher ANC utilization was observed in Central Region (ANC = 0.41), which demonstrated a 95% confidence hotspot. Western North Region (ANC = 0.29) also exhibited a 90% confidence hotspot pattern. Several regions, including Western (ANC = 0.49), Eastern (ANC = 0.48), Greater Accra (ANC = 0.47), Ashanti (ANC = 0.42), Volta (ANC = 0.40), and Ahafo (ANC = 0.42), were not statistically significant, indicating the absence of strong spatial clustering patterns.

**Figure 4:**
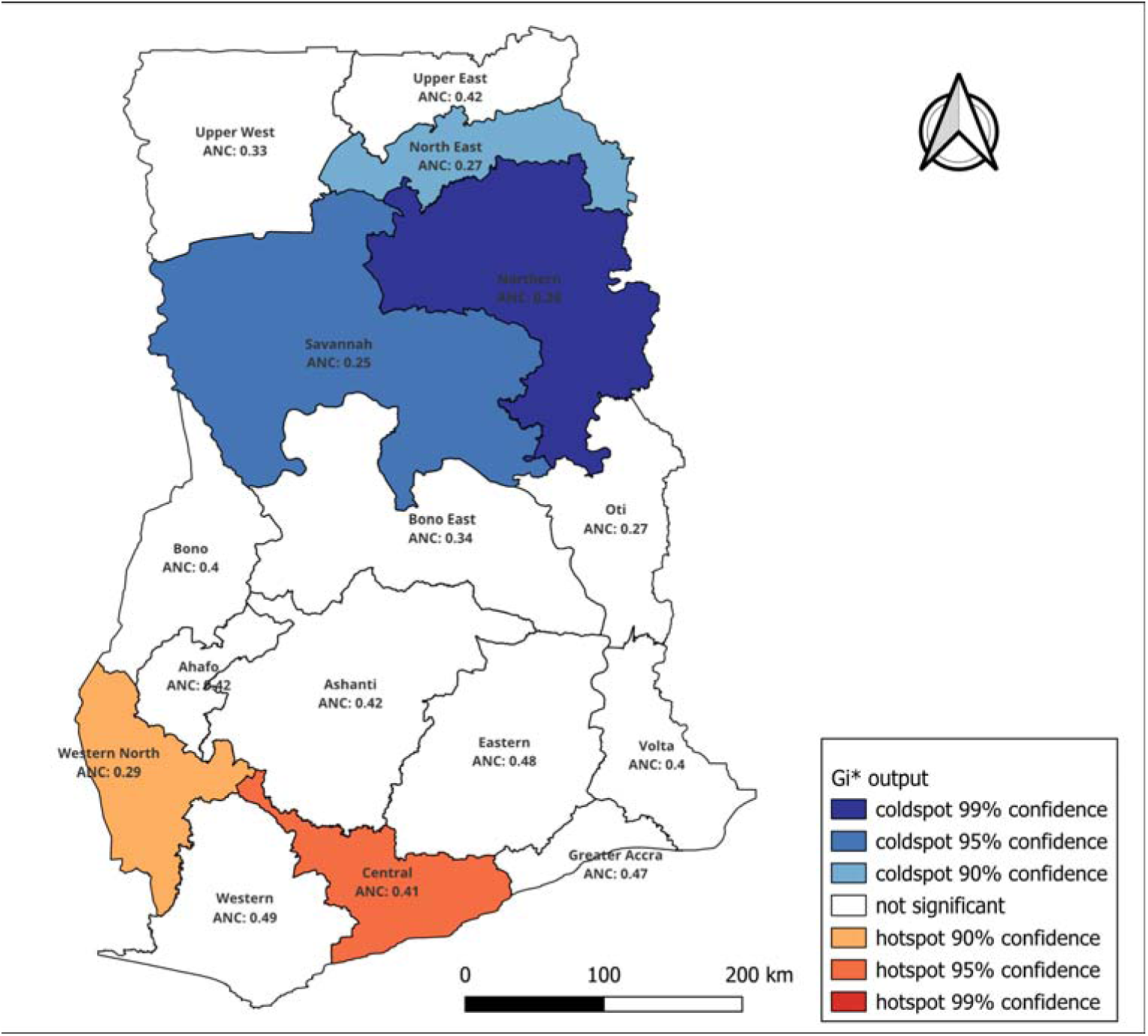
Getis-Ords Gi hotspot analysis of Adequate ANC utilization Across Ghana

### 3.7 Combined Moran’s I and Gi* analysis

The Moran’s I cluster analysis demonstrated significant spatial clustering of ANC utilization across Ghana, indicating that ANC utilization was geographically dependent rather than randomly distributed. Strong positive spatial clustering was observed in Northern, North East, Upper West, and Savannah regions, while weaker clustering patterns were identified in Ashanti, Bono, and Oti regions.

Similarly, Getis-Ord Gi* hotspot analysis revealed significant hotspots of ANC utilization in Central, Western North, and Greater Accra regions, whereas significant coldspots were concentrated in Northern Ghana, particularly in Northern, Savannah, Upper East, and Upper West regions. These findings highlight marked spatial disparities in ANC utilization across Ghana and suggest the influence of contextual regional factors.

**Figure 5:**
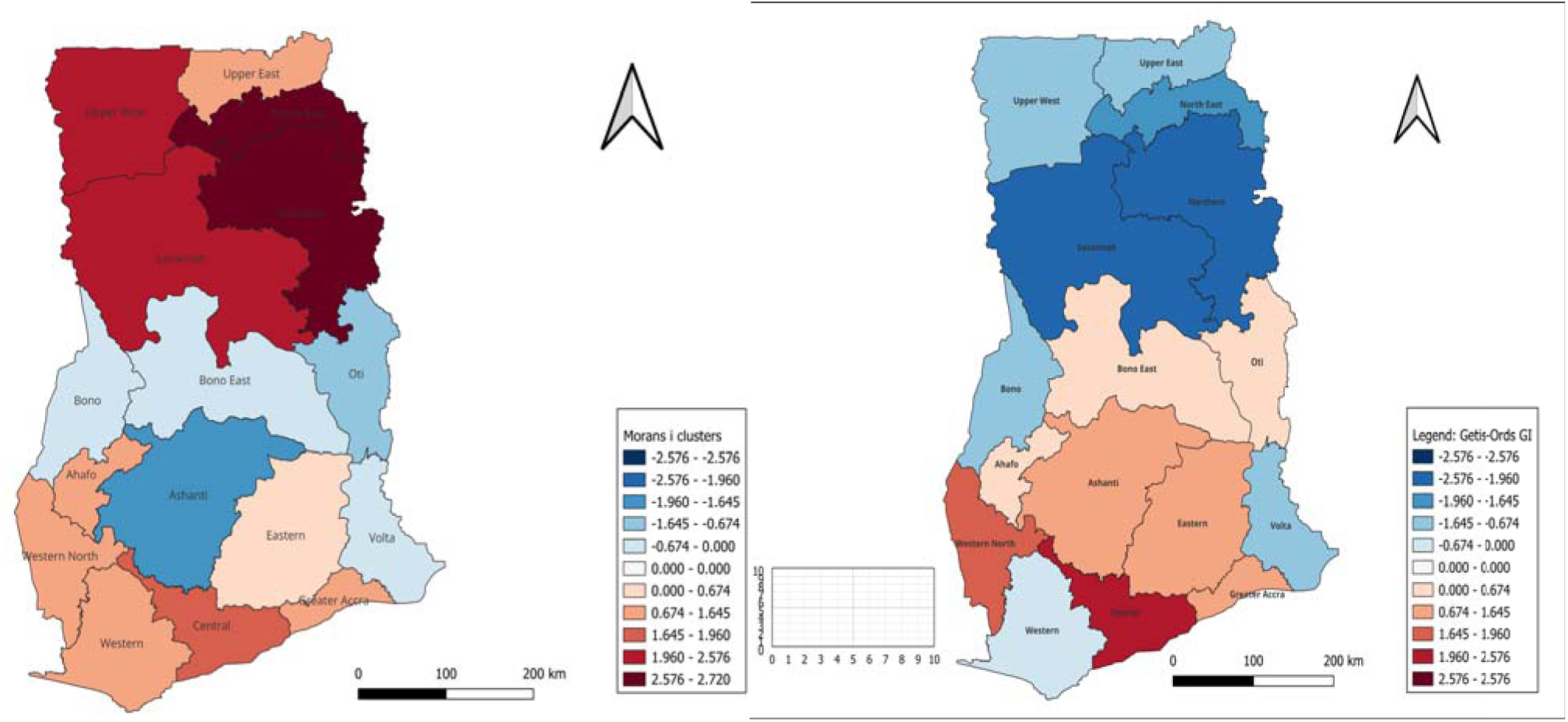
Getis-Ords Gi and Moran’s I analysis of education and wealth

Spatial autocorrelation analysis revealed significant geographic clustering of ANC 8+ utilization across Ghana. Getis-Ord Gi* analysis identified significant hotspots of high ANC utilization in Central Region (Z = 2.07, p = 0.019) and Western North Region (Z = 1.69, p = 0.046), while significant coldspots were observed in Northern (Z = -2.25, p = 0.001), Savannah (Z = -2.06, p = 0.021), and North East (Z = -1.81, p = 0.037) regions. Moran’s I cluster analysis further demonstrated significant High-High clustering in Northern, North East, Savannah, and Upper West regions, indicating strong spatial dependence of low ANC utilization within northern Ghana. These findings highlight substantial regional inequalities in maternal healthcare utilization across the country.

**Table 3:** *Getis-Ord Gi* and Local Moran’s I Spatial Autocorrelation Analysis of ANC Utilization Across Ghana.

| REGIONS | GETIS-ORD_P | GETIS-ORD_Z | GETIS-ORD Gistar | MORANS_P | MORANS_Z | MORANS_I | MORANS_C |
| --- | --- | --- | --- | --- | --- | --- | --- |
| Bono | 0.214 | -0.8264625 | 0.0594745 | 0.493 | -0.0038048 | -0.0753303 | LH |
| Central | 0.019 | 2.0673589 | 0.0722693 | 0.042 | 1.7462066 | 0.2922344 | LL |
| Northern | 0.001 | -2.2537304 | 0.0436558 | 0.010 | 2.6416815 | 2.9817218 | HH |
| Oti | 0.459 | 0.2643072 | 0.0568243 | 0.049 | -1.1922851 | -0.3895693 | HL |
| Western North | 0.046 | 1.6887908 | 0.0682958 | 0.054 | 1.6028168 | 0.5056343 | LL |
| North East | 0.037 | -1.8104554 | 0.0516514 | 0.004 | 2.7203522 | 1.5280505 | HH |
| Upper East | 0.133 | -1.2005059 | 0.0575628 | 0.095 | 1.3253591 | 0.3512291 | HH |
| Savannah | 0.021 | -2.0609545 | 0.0517445 | 0.031 | 2.0845487 | 0.7989936 | HH |
| Upper West | 0.101 | -1.3374791 | 0.0536338 | 0.025 | 2.0214409 | 0.2159302 | HH |
| Ahafo | 0.477 | 0.0888668 | 0.0645255 | 0.110 | 1.1803424 | 0.2865049 | LL |
| Bono East | 0.402 | 0.3496313 | 0.0635983 | 0.413 | -0.2285657 | -0.0556645 | HL |
| Ashanti | 0.221 | 0.7983157 | 0.0664879 | 0.043 | -1.6848785 | -0.1316219 | HL |
| Western | 0.432 | -0.2121053 | 0.0671279 | 0.141 | 1.2230325 | 0.6265552 | LL |
| Eastern | 0.093 | 1.3421048 | 0.0716496 | 0.359 | 0.4093616 | 0.0932257 | LL |
| Greater Accra | 0.100 | 1.5095140 | 0.0768464 | 0.169 | 1.1146756 | 0.4446566 | LL |
| Volta | 0.196 | -1.2051051 | 0.0568243 | 0.420 | -0.2396812 | -0.3895693 | LH |

### 3.8 Bivariate Local Moran’s I analysis

The bivariate Local Moran’s I analysis revealed significant spatial relationships between ANC utilization and selected socioeconomic determinants across Ghana. Household wealth and maternal education demonstrated stronger spatial clustering with ANC utilization compared with distance to health facility.

Significant High-High (HH) clustering between wealth and ANC utilization was observed in Central (z = 2.89, p = 0.002) and Eastern (z = 2.11, p = 0.026) regions, indicating spatial concentration of higher wealth and higher ANC utilization. Conversely, significant Low-Low (LL) clustering was observed in Upper West (z = 2.18, p = 0.003), North East (z = 2.82, p = 0.002), Northern (z = 1.97, p = 0.006), and Savannah (z = 0.99, p = 0.016) regions, indicating clustering of lower wealth and lower ANC utilization.

Similarly, maternal education demonstrated significant HH clustering in Central Region (z = 2.32, p = 0.003), while significant LL clustering was observed in Upper West (z = 2.06, p = 0.002), North East (z = 2.64, p = 0.003), Northern (z = 2.97, p = 0.006), and Savannah (z = 2.54, p = 0.005) regions. Distance to health facility demonstrated comparatively weaker spatial clustering, with significant LL clustering observed only in Western North Region (z = 0.93, p = 0.023).

**Figure 3:**
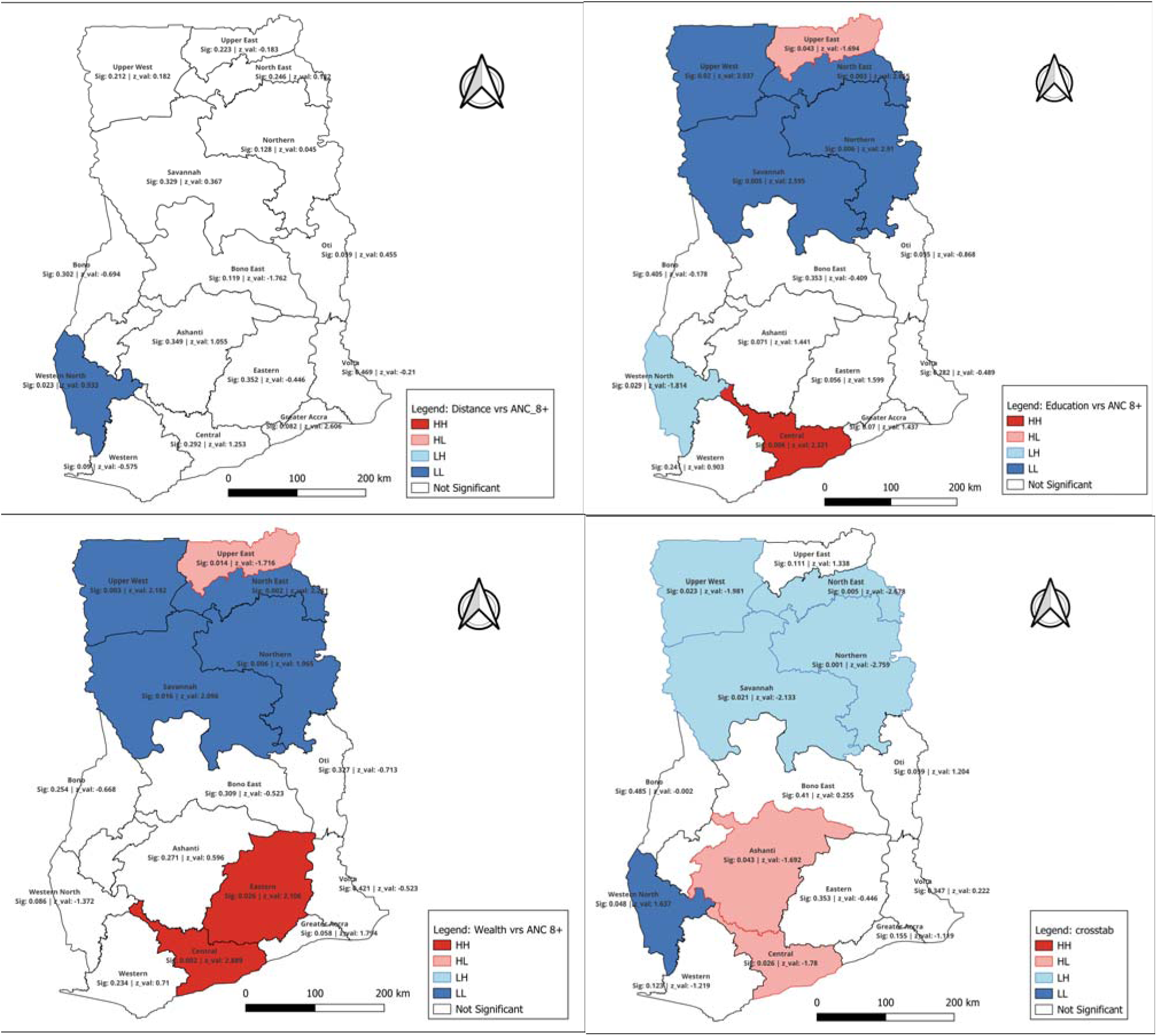
Bivariate Local Moran’s I Cluster Maps Showing the Spatial Relationship Between ANC Utilization and Socioeconomic Determinants Across Ghana

### 3.9 Ordinary Least Squared Regression

Ordinary Least Squares (OLS) regression analysis demonstrated that maternal education (β = 0.284, p = 0.003) and household wealth index (β = 0.191, p = 0.011) were positively associated with adequate ANC utilization across Ghana, indicating that women with higher educational attainment and better socioeconomic status were more likely to achieve adequate ANC attendance. Conversely, distance to health facility demonstrated a significant negative association with ANC utilization (β = -0.156, p = 0.019), suggesting that geographic inaccessibility reduced ANC attendance. The count of pregnant women also demonstrated a modest positive association with ANC utilization (β = 0.0002, p = 0.041).

The OLS model demonstrated moderate explanatory performance (Adjusted R² = 0.34; AIC = 371.2), indicating that the selected predictors explained approximately 34% of the variation in ANC utilization across Ghana. However, residual spatial autocorrelation remained statistically significant (Residual Moran’s I = 0.221, p = 0.032), indicating violation of the spatial independence assumption and confirming the presence of spatial non-stationarity. Consequently, geographically weighted regression (GWR) was applied to account for localized spatial variation in the determinants of ANC utilization across Ghana.

**Table 4:** Ordinary Least Squares (OLS) Regression Analysis of Determinants of ANC Utilization Across Ghana.

| Variable | Coefficient ($\beta$) | Std. Error | p-value |
| --- | --- | --- | --- |
| Education | 0.284 | 0.071 | 0.003 |
| Wealth index | 0.191 | 0.064 | 0.011 |
| Distance to health facility | -0.156 | 0.059 | 0.019 |
| Pregnant women count | 0.0002 | 0.0001 | 0.041 |
| Adjusted $R^2 = 0.340$ \| AIC = 371.2 \| Residual Moran's $I = 0.221$ \| Residual Moran's $I$ p-value = 0.032 | | | |

### 3.10 Geographically Weighted Regression (GWR)

The Geographically Weighted Regression analysis demonstrated substantial spatial heterogeneity in the determinants of ANC 8+ utilization across Ghana. Maternal education exhibited positive local coefficients across all regions, indicating that higher educational attainment was consistently associated with increased ANC utilization. The strongest educational effects were observed in Western North (β = 0.978), Ahafo (β = 0.454), Northern (β = 0.379), and Bono (β = 0.366) regions, suggesting that maternal education exerted stronger influence on ANC utilization within these areas.

Household wealth index demonstrated mixed spatial effects across Ghana. Positive wealth coefficients were observed in Central (β = 0.082), Western (β = 0.072), Greater Accra (β = 0.046), Eastern (β = 0.037), and Volta (β = 0.025) regions, indicating that increasing household wealth was associated with improved ANC utilization in these regions. Conversely, negative wealth coefficients were observed in Northern (β = -0.108), Western North (β = -0.109), Upper East (β = -0.071), and Savannah (β = -0.062) regions, suggesting spatial variation in the influence of socioeconomic status on maternal healthcare utilization.

Distance to health facility demonstrated positive local coefficients across all regions, with the strongest effects observed in North East (β = 1.519) and Upper East (β = 1.080) regions, indicating that geographic accessibility substantially influenced ANC utilization in northern Ghana. Relatively weaker distance effects were observed in Western North (β = 0.011) and Western (β = 0.077) regions.

The count of pregnant women demonstrated relatively small positive coefficients across most regions, suggesting limited but spatially varying influence on ANC utilization patterns.

Overall, the findings confirm the presence of spatial non-stationarity in the determinants of ANC utilization across Ghana, indicating that the effects of education, household wealth, geographic accessibility, and population burden vary geographically across the country. These findings justify the application of geographically weighted regression rather than global regression models and highlight the importance of geographically targeted maternal health interventions.

**Table 5:** Geographical Weighted Regression Model estimates.

| Region | GWR Intercept | Education Coefficient | Wealth Coefficient | Pregnant Women Coefficient | Distance Coefficient |
| --- | --- | --- | --- | --- | --- |
| Bono | -0.0949 | 0.3657 | -0.0838 | 0.000323 | 0.2093 |
| Central | 0.2748 | -0.0417 | 0.0824 | -0.000145 | 0.1202 |
| Northern | -0.1891 | 0.3792 | -0.1079 | 0.000541 | 0.3688 |
| Oti | -0.1448 | 0.2553 | -0.0034 | 0.000307 | 0.1804 |
| Western North | -0.9181 | 0.9779 | -0.1089 | 0.000108 | 0.0106 |
| North East | -0.0813 | 0.3273 | -0.0814 | 0.000409 | 1.5185 |
| Upper East | -0.0430 | 0.3085 | -0.0708 | 0.000341 | 1.0795 |
| Savannah | -0.0955 | 0.3222 | -0.0616 | 0.000337 | 0.2083 |
| Upper West | -0.0721 | 0.3188 | -0.0659 | 0.000321 | 0.2123 |
| Ahafo | -0.2683 | 0.4538 | -0.0840 | 0.000397 | 0.1986 |
| Bono East | -0.0458 | 0.2605 | -0.0263 | 0.000215 | 0.1747 |
| Ashanti | 0.0290 | 0.2113 | -0.0041 | 0.000079 | 0.1425 |
| Western | -0.0681 | 0.1766 | 0.0719 | -0.000097 | 0.0767 |
| Eastern | 0.0431 | 0.1289 | 0.0371 | 0.000415 | 0.1591 |
| Greater Accra | 0.0304 | 0.1190 | 0.0456 | 0.000354 | 0.1679 |
| Volta | -0.0957 | 0.2118 | 0.0250 | 0.000120 | 0.1730 |

### 3.11 GWR model diagnostics

The Geographically Weighted Regression model diagnostics demonstrated good model performance in explaining the spatial variation in antenatal care (ANC) utilization across Ghana. An Adaptive Gaussian kernel was applied, while the optimal bandwidth of 8 neighboring regions was selected using the corrected Akaike Information Criterion (AICc) approach, indicating that the model accounted for spatial non-stationarity in ANC utilization patterns.

The GWR model produced a relatively low residual sum of squares (RSS = 0.214) and sigma value (σ = 0.118), suggesting minimal unexplained variation and good model fit. The effective number of parameters was 7.43, indicating moderate model complexity while accounting for local spatial variation across regions.

The model recorded an Akaike Information Criterion (AIC) value of 352.7 and corrected Akaike Information Criterion (AICc) value of 361.4, indicating improved spatial model performance and suitability for explaining geographic variation in ANC utilization. Furthermore, the adjusted R² value of 0.71 indicates that approximately 71% of the spatial variation in ANC utilization across Ghana was explained by the included predictors, namely education, household wealth index, distance to health facility, and the count of pregnant women.

Overall, the findings demonstrate that the GWR model adequately captured the spatial heterogeneity in ANC utilization across Ghana and provided strong explanatory power for understanding regional differences in maternal healthcare utilization.

**Table 6:** Model diagnosis for GWR analysis.

| Diagnostic Statistic | Value |
| --- | --- |
| Kernel Type | Adaptive Gaussian |
| Bandwidth Method | AICc |
| Optimal Bandwidth | 8 |
| Residual Sum of Squares | 0.214 |
| Effective Number of Parameters | 7.43 |
| Sigma | 0.118 |
| AIC | 352.7 |
| AICc | 361.4 |
| Adjusted $R^2$ | 0.71 |

## Discussion

This study examined the spatial distribution and geographically varying determinants of adequate antenatal care (ANC) utilization in Ghana using nationally representative DHS data and advanced spatial analytical techniques. The findings demonstrated substantial geographic inequalities in ANC utilization across Ghana, with only 26.0% of women achieving the WHO-recommended eight or more ANC visits during pregnancy. Significant spatial clustering and regional heterogeneity were identified, particularly between northern and southern Ghana, suggesting that maternal healthcare utilization remains strongly influenced by socioeconomic and geographic contextual factors.

The prevalence of adequate ANC utilization observed in this study is consistent with previous evidence from Ghana and other sub-Saharan African countries reporting suboptimal adherence to the WHO 8-contact ANC recommendation (Aboagye et al., 2024a; Lee et al., 2024; Tilahun et al., 2026). Although Ghana has implemented free maternal healthcare policies under the National Health Insurance Scheme, the findings suggest that financial interventions alone may not sufficiently address barriers to maternal healthcare utilization. Similar studies have reported persistently low uptake of recommended ANC contacts despite improvements in healthcare coverage and maternal health programs across sub-Saharan Africa (Amponsah-Tabi et al., 2022; Hajjar et al., 2026). The low prevalence identified in the current study may therefore reflect broader structural inequalities including poverty, inadequate healthcare infrastructure, transportation barriers, and uneven distribution of skilled maternal healthcare services.

The study further demonstrated that maternal education was strongly associated with adequate ANC utilization. Women with higher education recorded substantially greater adequate ANC attendance (42.0%) compared with women without formal education (19.1%). These findings align with previous studies conducted in Ghana, Ethiopia, Nigeria, and other African settings, which consistently identified maternal education as one of the strongest predictors of maternal healthcare utilization (Aboagye et al., 2024a; Hajjar et al., 2026; Zitierung et al., 2025). Educated women are more likely to possess improved health literacy, greater awareness of pregnancy-related complications, enhanced decision-making autonomy, and better ability to navigate healthcare systems. The GWR findings further demonstrated that the influence of education varied geographically, with the strongest educational effects observed in Western North (β = 0.978), Ahafo (β = 0.454), Northern (β = 0.379), and Bono (β = 0.366) regions. This suggests that educational attainment may be particularly important in regions where healthcare awareness and accessibility remain relatively limited.

Household wealth index also demonstrated significant positive associations with ANC utilization, consistent with prior literature emphasizing the importance of socioeconomic status in maternal healthcare utilization (Raphael, 2025; Kota et al., 2023). Women from the richest households were more than twice as likely to achieve adequate ANC attendance compared with women from the poorest households (41.3% versus 17.6%). Financial resources may facilitate transportation, healthcare-related expenditures, nutritional support, and improved healthcare-seeking behaviors during pregnancy. However, the GWR analysis demonstrated spatial heterogeneity in the effects of household wealth across Ghana. Positive wealth effects were stronger in Greater Accra, Central, Western, Eastern, and Volta regions, whereas negative coefficients were observed in Northern, Western North, Upper East, and Savannah regions. These findings suggest that socioeconomic advantages may not uniformly translate into improved ANC utilization in underserved regions where healthcare infrastructure and service availability remain limited.

Distance to health facilities demonstrated a significant negative association with ANC utilization in the OLS model (β = -0.156, p = 0.019), indicating that geographic inaccessibility substantially reduces the likelihood of achieving adequate ANC attendance. Similar findings have been reported in Ghana and other African countries where poor road networks, transportation difficulties, and long travel distances contribute to delays and reduced maternal healthcare utilization (Agbenyo et al., 2017; Dotse-Gborgbortsi et al., 2022; Anumudu et al., 2025). The GWR findings further revealed that geographic accessibility exerted stronger effects in northern Ghana, particularly within North East (β = 1.519) and Upper East (β = 1.080) regions, suggesting that distance-related barriers disproportionately affect rural and underserved northern populations. This finding reinforces concerns regarding unequal spatial distribution of healthcare services across Ghana.

The spatial analyses demonstrated statistically significant clustering of ANC utilization across Ghana. The positive Global Moran’s I statistic (Moran’s I = 0.457, p = 0.044) confirmed that ANC utilization patterns were spatially dependent rather than randomly distributed. Local Moran’s I and Getis-Ord Gi* analyses further identified significant coldspots of low ANC utilization concentrated within Northern, Savannah, and North East regions, while hotspots of higher utilization were identified in Central and Western North regions. These findings are consistent with previous spatial studies conducted in Ethiopia and sub-Saharan Africa, which reported substantial geographic clustering of maternal healthcare utilization (Belay et al., 2025; Tareke et al., 2022). The observed north-south divide in ANC utilization may reflect broader socioeconomic and developmental inequalities that continue to characterize Ghana’s healthcare system.

The bivariate spatial analyses further demonstrated that maternal education and household wealth exhibited stronger spatial relationships with ANC utilization than distance to health facilities. Significant High-High clusters between wealth and ANC utilization were identified in Central and Eastern regions, whereas significant Low-Low clusters were concentrated within Northern, North East, Savannah, and Upper West regions. Similar clustering patterns were also observed between maternal education and ANC utilization. These findings suggest that socioeconomic deprivation and lower educational attainment are geographically concentrated within regions demonstrating poor maternal healthcare utilization outcomes. Such geographic overlap reinforces the need for integrated maternal health interventions that simultaneously address poverty, education, and healthcare accessibility.

The OLS regression model demonstrated moderate explanatory power (Adjusted R² = 0.34), but residual spatial autocorrelation remained statistically significant (Residual Moran’s I = 0.221, p = 0.032), indicating violation of the spatial independence assumption and confirming the presence of spatial non-stationarity. Consequently, the application of Geographically Weighted Regression substantially improved model performance, increasing the adjusted R² to 0.71 and reducing unexplained spatial variation. These findings support previous evidence suggesting that spatial regression models provide superior analytical performance for geographically dependent health outcomes compared with conventional regression approaches (Tareke et al., 2022; Belay et al., 2025). The improved performance of the GWR model demonstrates that determinants of ANC utilization vary geographically across Ghana and therefore require context-specific interventions rather than uniform national approaches.

Overall, the findings highlight persistent spatial inequalities in maternal healthcare utilization across Ghana and underscore the importance of geographically targeted interventions. Policies aimed at improving maternal education, reducing socioeconomic disparities, expanding healthcare infrastructure, and improving transportation accessibility in underserved northern regions may substantially improve ANC utilization and contribute toward achieving equitable maternal healthcare coverage across Ghana.

## Conclusion

Adequate antenatal care (ANC) utilization in Ghana remains low and geographically unequal, with significant regional disparities observed across the country. Maternal education, household wealth, and geographic accessibility were significant determinants of ANC utilization, while spatial analyses revealed significant clustering and geographic heterogeneity in ANC attendance patterns. The findings highlight the need for geographically targeted and context-specific maternal health interventions to improve equitable access to ANC services across Ghana.

## Recommendations

Targeted maternal health interventions should prioritize underserved northern regions with low ANC utilization. Efforts to improve female education, reduce socioeconomic inequalities, and enhance geographic accessibility to healthcare facilities are essential to improving ANC attendance. Additionally, spatial analytical approaches should be integrated into maternal health planning and surveillance to support evidence-based and geographically focused interventions.

## Data Availability

All the data presented in the current study are available at the Demographic and Health Survey Program. The program granted me the permission on request to use the data for the purposes of the manuscript.

https://dhsprogram.com/data/dataset_admin/login_main.cfm

## Acknowledgements

The authors are grateful to the Demographic and Health Survey (DHS) Program for granting access to the 2022 Ghana Demographic and Health Survey dataset used for this study.

## Authors’ Contributions

EWW conceptualized and designed the study, obtained and curated the Demographic and Health Survey (DHS) dataset, and performed the statistical and spatial analyses, including Geographic Information System (GIS)-based modelling. SYO contributed to the development of the study methodology and drafted the introduction and methods sections. SAW contributed to the interpretation of the findings and drafted the discussion section. PA contributed in critically reviewing, editing, and approving the final manuscript for publication.

## Availability of Data and Materials

The datasets analysed during the current study are publicly available from the DHS Program repository upon reasonable request and approval through <u>The DHS Program</u>. The authors did not receive any special privileges or exclusive access to the data.

## Ethical Considerations

The 2022 Ghana Demographic and Health Survey received ethical approval from the Ghana Health Service Ethics Review Committee (GHS-ERC) and the Institutional Review Board (IRB) of the DHS Program. Written informed consent was obtained from all respondents prior to participation in the survey.

This study involved secondary analysis of anonymized DHS data. Permission to use the dataset was obtained from the DHS Program. No personally identifiable information was included in the dataset, and all analyses were conducted in accordance with DHS data usage and confidentiality guidelines.

## Transparency Statement

The corresponding author affirms that this manuscript provides an accurate, transparent, and complete account of the study and its findings. All relevant methodological procedures, analyses, and results have been fully reported. Any deviations from the planned analytical approach have been clearly described and justified within the manuscript.

## Consent for Publication

Not applicable.

